# Small Fiber Neuropathy after COVID-19: A Key to Long COVID

**DOI:** 10.1101/2023.11.07.23297764

**Authors:** Lindsay S. McAlpine, Adeel S. Zubair, Phillip Joseph, Serena S. Spudich

## Abstract

**Objectives:** Report a case series of new onset small fiber neuropathy (SFN) after COVID-19 treated with intravenous immunoglobulin (IVIG). SFN is a critical objective finding in long COVID and amenable to treatment.

**Methods:** A retrospective chart review was conducted on patients seen in the NeuroCOVID Clinic at Yale who developed new-onset SFN after a documented COVID-19 illness. We documented demographics, symptoms, treatments, diagnostics, and clinical response to treatment.

**Results:** Sixteen patients were diagnosed with length dependent or independent SFN on skin biopsy (median age 47, 75% female, 75% Caucasian). Among the nine patients tested for autoantibodies, six were positive for either trisulfated heparin disaccharide (TS-HDS) or fibroblast growth factor receptor 3 (FGFR3). Eight patients underwent treatment with IVIG and experience significant clinical improvement in their neuropathic symptoms. 92% of patients reported post-exertional malaise characteristic of myalgic encephalomyelitis/chronic fatigue syndrome (ME/CFS) and six patients underwent invasive cardiopulmonary exercise testing (iCPET), which demonstrated neurovascular dysregulation and dysautonomia consistent with ME/CFS.

**Discussion:** Here we present preliminary evidence that SFN is responsive to treatment with IVIG and linked with neurovascular dysregulation and dysautonomia. A larger clinical trial is indicated to further demonstrate the clinical utility of IVIG in treating post-infectious small fiber neuropathy.

## Background

Patients with long COVID-19 frequently present with neuropathic symptoms including numbness, paresthesias, allodynia, and autonomic dysfunction consistent with small fiber neuropathy (SFN). Preliminary evidence suggests small fiber neuropathy may be a key pathologic finding in long COVID.^1-4^ Many patients with long COVID also have overlapping symptomatology with Myalgic Encephalomyelitis/Chronic Fatigue Syndrome (ME/CFS) and/or Postural Orthostatic Tachycardia Syndrome (POTS). Prior to the pandemic, ME/CFS and POTS have been linked to SFN.^5-9^ It is hypothesized that the inflammatory immune response during a viral illness may lead to immune dysregulation and damage to small fiber nerves.^1^ We report 16 patients who developed post-COVID SFN and their outcomes with immunotherapy.

## Methods

A retrospective chart review was conducted on patients seen in the NeuroCOVID Clinic at Yale who developed new-onset small fiber neuropathy after a documented COVID-19 illness. We documented demographics, symptoms, treatments, diagnostics, and clinical response to treatment. The sensory neuropathy autoantibody testing was completed by the Neuromuscular Clinical Laboratory at Washington University at St. Louis. Inclusion criteria included diagnosis of long COVID based on the World Health Organization (WHO) definition, no prior neuropathy diagnosis, and positive punch skin biopsy (defined as reduced Intraepidermal Nerve Fiber Density (IENFD) on bright field immunohistochemistry by Corinthian Reference Lab). Inclusion criteria also included negative electrodiagnostic testing and negative laboratory testing for other causes of neuropathy. Exclusion criteria included symptom onset after the vaccine and prior diagnosis of neuropathy from any cause. We received IRB approval to conduct this retrospective study was received and participant consent was waived.

## Results

Sixteen patients were diagnosed with small fiber neuropathy on skin biopsy. The median age was 47 (IQR 40 – 58), 75% were female (n = 12), and 75% were Caucasian (n = 12) (**Table 1**). 38% of patients were hospitalized during their acute COVID-19 illness on the medicine floor, but none required ICU level care. The majority of patients had a mild COVID-19 illness. The onset of neuropathic symptoms, including numbness, paresthesias, and allodynia, was a median of 2.5 weeks (IQR 2 – 3.75) after the first day of the COVID-19 illness. No patient had a prior diagnosis of neuropathy and no patient had neuropathic symptoms prior to their COVID-19 illness, except for one who reported intermittent numbness on the plantar aspect of their feet. All patients demonstrated reduced IENFD on skin biopsy, the majority of which had length dependent SFN (56%). The presence of SFN was associated with a constellation of dysautonomia symptoms, including orthostasis (69%), altered sweating (77%), labile heart rate (85%), and post-exertional malaise (92%). Nine patients underwent sensory neuropathy autoantibody testing: 3 were positive for trisulfated heparin disaccharide (TS-HDS) and 3 were positive for fibroblast growth factor receptor 3 (FGFR3).

**Table 1:** Demographics and SFN characteristics in our case series cohort.

| **Median (IQR) \| n (%)** |  |  |
| --- | --- | --- |
| **n = 16** |  |  |
| **Age** | 47 (40 – 58) |  |
| **Female Gender** | 75% |  |
| **Race/Ethnicity** |  |  |
| *Caucasian* | 12 (75%) |  |
| *Black* | 3 (19%) |  |
| *Hispanic* | 1 (6%) |  |
| **Hospitalized for COVID-19** | 6 (38%) |  |
| **Symptom Onset After COVID-19 (weeks)** | 2.5 (2 – 3.75) |  |
| **Pre-Existing Neuropathic Symptoms** | 1 (6%) |  |
| **Diabetes** | 3 (19%) |  |
| **Pre-Diabetes** | 2 (13%) |  |
| **Type of SFN** |  |  |
| *Length Dependent* | 10 (56%) |  |
| *Non-Length Dependent* | 6 (38%) |  |
| **Autonomic Symptoms** |  |  |
| *Orthostasis* | 10 (69%) |  |
| *Altered Sweating* | 10 (77%) |  |
| *Heart Rate Lability* | 11 (85%) |  |
| *Post-Exertional Malaise* | 12 (92%) |  |
| *GI/Bladder Dysfunction* | 2 (15%) |  |
| **Antibody Testing** | **n = 9** | **Median (range)** |
| *FGFR3* | 3 (33%) | 6,800 (6,000-9,000) |
| *TS-HDS* | 3 (33%) | 23,000 (14,000-57,000) |

Eight patients received intravenous immunoglobulin (IVIG) for a median of 9.5 months (IQR 3 – 18.5) (**Table 2**). The majority of patients (n=5; 63%) experienced resolution of their clinical neuropathic symptoms. The remaining 37% (n=3) of patients a significant improvement in the extent and intensity of their symptoms. Notably, the patients without complete resolution had a diagnosis of diabetes or pre-diabetes.

**Table 2:** Clinical response to immunotherapy in a subset of patients who underwent treatment with IVIG.

| Median (IQR) \| n (%) |  |
| --- | --- |
| n = 8 |  |
| **Onset of symptoms after COVID-19 infection onset (weeks)** | 3 (2 – 3) |
| **Length of IVIG Treatment (months)** | 9.5 (3 – 18.5) |
| **Response to IVIG** |  |
| *Resolved* | 5 (63%) |
| *Improvement* | 3 (37%) |
| **Distribution** |  |
| *Extremities* | 8 (100%) |
| *Trunk* | 1 (13%) |
| **Symptoms** |  |
| *Allodynia* | 2 (25%) |
| *Numbness* | 6 (75%) |
| *Tingling/Burning* | 8 (100%) |
| *Poor Proprioception* | 4 (50%) |

Seven patients (44%) who presented with post-exertional malaise and autonomic symptoms underwent a clinical invasive cardiopulmonary exercise test (iCPET). All 7 patients demonstrated evidence of neurovascular dysregulation and dysautonomia consistent with a diagnosis of ME/CFS. Findings included depressed aerobic capacity, elevated peak mixed venous oxygen saturation, depressed arterial-venous oxygen content difference, and low biventricular filling pressures.

## Discussion

SFN should be considered in the differential for patients who present with neuropathic symptoms and autonomic dysfunction after COVID-19. SFN neuropathic symptoms commonly include distal, symmetric burning pain, allodynia, impaired temperature sensation, paresthesias, and numbness. SFN autonomic symptoms include abnormal sweating, skin discoloration, cool extremities, dysautonomia, and bladder and bowel issues. Electrodiagnostic testing is negative in SFN, but a skin punch biopsy, which looks for a reduction in small nerve fiber density, will support the diagnosis. A skin punch biopsy is not specific for an etiology of SFN. Autoantibody testing may include TS-HDS, FGFR3, and Histone H3, but their clinical relevance is still under debate. TS-HDS and FGFR3 autoantibodies have been linked with SFN and dysautonomia previously.^10^ The most common causes of SFN should also be evaluated for, including diabetes, HIV, autoimmune diseases such as Sjogrens, SLE, Celiac disease, and sarcoidosis, and toxic exposures such as alcohol and chemotherapy.

Given the time of onset within 6 weeks of an acute illness and response to immunotherapy, we propose SFN after COVID-19 has an underlying autoimmune etiology. The pathophysiology of small fiber neuropathy may include antibody mediated damage, molecular mimicry, and/or smoldering inflammation.^11-14^ SFN has been linked to both ME/CFS and POTS and we provide further evidence here (**Figure 1**).^5,7-9,15^ ME/CFS is characterized by post-exertional malaise not alleviated by rest leading to significant functional impairment. It is often accompanied by cognitive impairment, orthostatic intolerance, and poor sleep. It is hypothesized that neurovascular dysregulation from damaged small nerve fibers innervating the microvasculature causes inappropriate dilation and shunting of oxygenated blood away from exercising muscles, which leads to exercise intolerance.^6^ There are limitations in our analysis and potential bias in this cohort given they were all selected from a NeuroCOVID Clinic. Further research is needed to better understand the pathophysiology of damage to small fiber nerves after COVID-19 and the connection to ME/CFS. Additionally, a larger clinical trial is indicated to further demonstrate the clinical utility of IVIG in treating post-infectious small fiber neuropathy.

**Figure 1.**
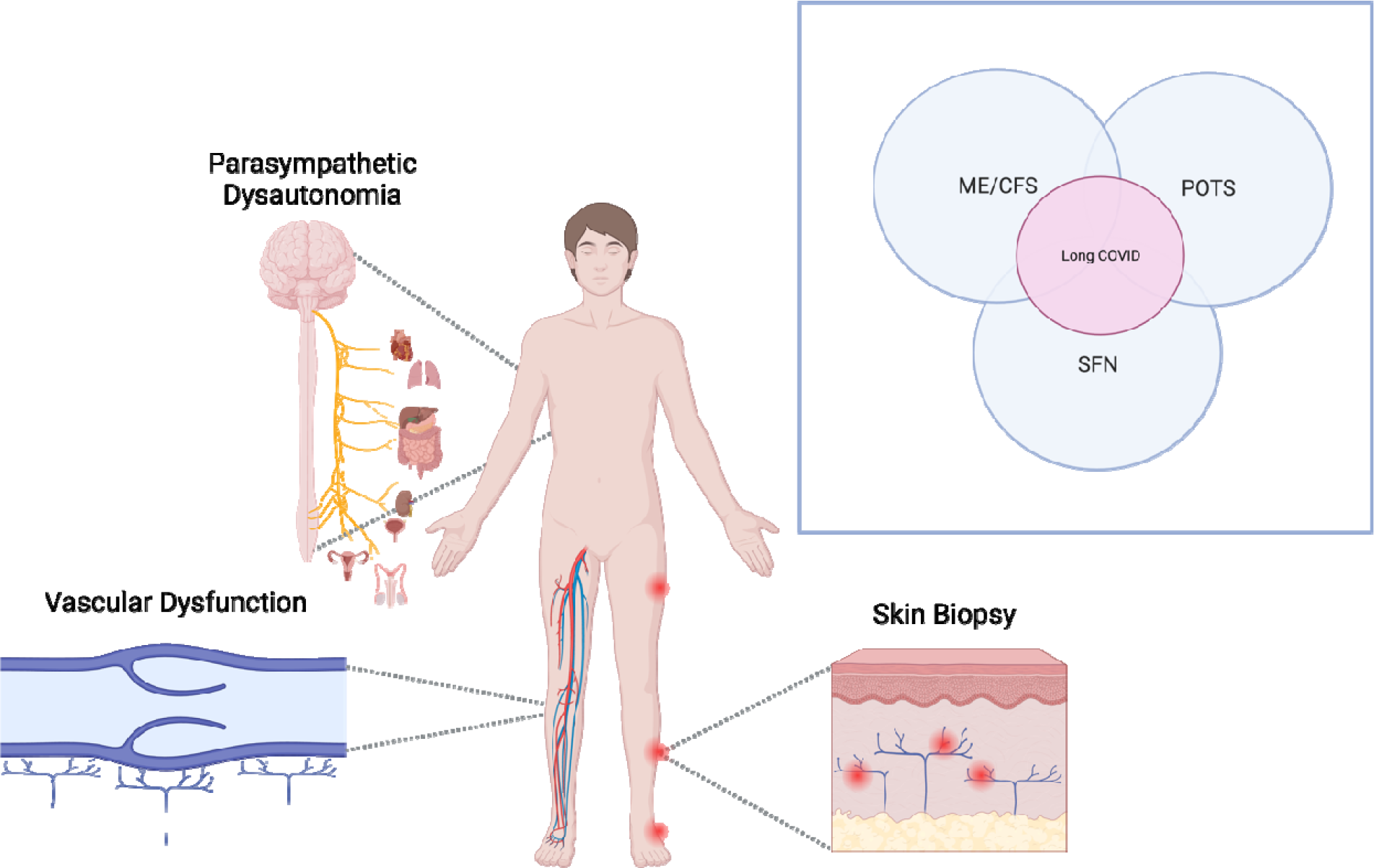
Long COVID and Small Fiber Neuropathy. Small fiber neuropathy can cause parasympathetic and microvascular dysfunction. Both ME/CFS and POTS have been linked to dysautonomia. Individuals with long COVID may experience one or more of these conditions. SFN is typically diagnosed with a skin biopsy evaluating the nerve fiber density.

## Data Availability

All data produced in the present work are contained in the manuscript

